# Single-nucleus transcriptomic analysis of pediatric pancreas reveals cellular heterogeneity and early neoplasia signatures during chronic pancreatitis

**DOI:** 10.64898/2026.07.01.26357053

**Authors:** Faizan Ahmed, Xiangfei Xie, Ajay Dixit, Maria E. Moreno-Fernandez, Erika Hissong Patel, Juan Gurria, Karmah Khoury, Phoebe Christian, Rita Bottino, Rathnakumar Kumaragurubaran, David Adeleke, Clive H. Wasserfall, Yunguan Wang, Maisam Abu-El-Haija

## Abstract

**Background:** Pediatric chronic pancreatitis (CP) carries an elevated lifetime risk of pancreatic ductal adenocarcinoma (PDAC), yet the cellular and molecular mechanisms driving disease progression and early neoplastic transformation remain undefined.

**Methods:** We performed single-nucleus RNA sequencing (snRNA-seq) on pancreatic tissue from 15 pediatric CP individuals and 6 healthy controls (HC). Findings were integrated with peripheral blood flow cytometry immunophenotyping of 8 CP and 7 HC individuals and validated by histopathological assessment.

**Findings:** We identified 15 distinct cell populations and profound cellular remodeling in CP, including a 46% reduction in acinar cells and emergence of inflammatory fibroblasts as the dominant stromal population. Acinar-to-ductal metaplasia (ADM) and pancreatic intraepithelial neoplasia (PanIN) populations bearing early PDAC-associated transcriptional signatures were detected in most CP samples. Cell-cell interaction analysis revealed that 68% of CP-specific ligand-receptor interactions converged on ADM and PanIN populations via ECM-integrin and inflammatory pathways. Peripheral blood flow cytometry demonstrated concordant systemic immune activation, including elevated monocyte CCR2 and CD80, increased CD69 on T cells, and upregulated RORγt in regulatory T cells.

**Interpretation:** This atlas defines the cellular landscape and intercellular signaling networks underlying pediatric CP, identifying inflammatory fibroblasts and early neoplastic cell states as central features. These findings provide a molecular foundation for understanding cancer risk in pediatric CP and provide a resource to prioritize studies into potential therapeutic targets and biomarkers.

**Funding:** This work was supported by the Network for Pancreatic Organ donors with Diabetes (nPOD) and The Leona M. & Harry B. Helmsley Charitable Trust.

**Research in context:** *Evidence before this study:* Chronic pancreatitis (CP) is a progressive inflammatory condition marked by irreversible structural alterations, fibrosis, and the deterioration of pancreatic function, leading to considerable morbidity and a diminished quality of life. Prior studies have characterized immune infiltration patterns and fibroblast heterogeneity in adult CP and pancreatic cancer using single-cell approaches, and spatial transcriptomics have been applied to adult pancreatic tissues. A single-nucleus atlas of the normal human pancreas from neonatal and adult donors provided a cellular reference framework. However, no study had applied single-cell or single-nucleus transcriptomics to pediatric CP tissue. The cellular composition, immune activation states, fibroblast diversity, and presence of early neoplastic signatures in the pediatric CP pancreas were entirely uncharacterized, leaving a critical gap in understanding disease mechanisms in this population with distinct genetic etiology and elevated lifetime cancer risk.

*Added value of this study:* We generated the first single-nucleus transcriptomic atlas of pediatric CP by profiling nuclei from pancreatic tissue of 15 children with CP and 6 healthy controls. We identified significantly distinct cell populations in CP compared to HC and uncovered marked cellular remodeling, including significant reduction in acinar cells, major immune cell expansion, and the emergence of inflammatory fibroblasts. Critically, we detected acinar-to-ductal metaplasia and pancreatic intraepithelial neoplasia populations bearing PDAC-associated transcriptional signatures in most CP samples, providing the first molecular evidence of early neoplastic changes in a pediatric context. Cell-cell interaction analysis revealed that inflammatory fibroblasts preferentially signal to precancerous populations via ECM-integrin and metabolic pathways. Complementary peripheral blood flow cytometry demonstrated concordant systemic immune activation, linking tissue-level inflammation to circulating immune dysregulation.

*Implications of all the available evidence:* Taken together, the available evidence establishes that pediatric CP is characterized by profound cellular remodeling, inflammatory fibroblast-driven signaling to precancerous cell populations, and early molecular features of neoplastic transformation that may precede histologically identifiable lesions by years. The detection of ADM and early PanIN signatures in children raises an important question about whether current cancer surveillance guidelines, which recommend screening from age 40 to 50 years in hereditary pancreatitis, should be reassessed for pediatric-onset disease. Circulating immune markers, including monocyte CCR2, CD80, and T-cell CD69, represent candidate biomarkers for disease monitoring. These findings may inform the design of host-directed therapies aimed at modulating immune-stromal interactions to attenuate fibrosis and reduce long-term cancer risk. The cellular programs identified here may also be relevant to other forms of pediatric inflammatory pancreatic diseases.

## Introduction

Chronic pancreatitis (CP) is a progressive inflammatory disorder characterized by irreversible structural changes, fibrosis, and loss of pancreatic function, resulting in significant morbidity and reduced quality of life^1,2^. While adult CP has been extensively studied, CP in pediatric populations remains poorly characterized at the cellular and molecular levels, despite its distinct etiology, progression, and long-term consequences^3,4^. Notably, pediatric CP lacks comprehensive characterization, representing a critical unmet need to understand its unique pathogenesis, which differs in their risk factors and disease progression from that of adult CP.^5^

The incidence of CP in children is rising globally, with an estimated two cases per 100,000 children annually^6,7^. Unlike adult CP, which is predominantly linked to environmental factors such as alcohol consumption and smoking, pediatric CP is more commonly associated with genetic mutations, anatomical anomalies, and systemic diseases^8,9^. Genetic variants in *PRSS1*, *SPINK1*, *CFTR*, and other genes are present in up to 70% of children with CP, underscoring the major role of hereditary factors in disease development^10,11^. Hereditary pancreatitis markedly increases the risk of pancreatic ductal adenocarcinoma (PDAC), with affected individuals experiencing up to a 53-fold increased risk compared to the general population, a phenomenon only described in adulthood age ranges^12^. Moreover, CP increases the risk of pancreatic cancer by nearly eight-fold within five years of diagnosis, highlighting the importance of long-term surveillance in individuals with CP^13^. Despite this, the cellular landscape and precancerous changes in pediatric CP remain unexplored, limiting our understanding of early neoplastic transformation and impeding therapeutic development.

Recent advances in single-cell and single-nucleus RNA sequencing (snRNA-seq) have transformed our ability to resolve tissue heterogeneity and cellular interactions in complex diseases at unprecedented resolution^14,15^. These approaches have been successfully applied to adult pancreatic disorders, including PDAC, revealing unexpected cellular diversity and novel pathogenic mechanisms^16,17^. For instance, pancreas tissue-resident macrophages activation during pancreatitis has been shown to drive the emergence of fibroblast subpopulations with divergent roles in extracellular matrix remodeling, implicating stromal heterogeneity as a key contributor to cancer pathogenesis^18^. Single cell profiling of experimental pancreatitis models has further revealed complex immune cell infiltration patterns, highlighting the interplay between innate and adaptive immunity during pancreatic inflammation^18,19^. A recent snRNA-seq atlas of the human pancreas, profiling nuclei from neonatal and adult donors, identified multiple acinar and ductal subtypes and established a reference framework of pancreatic cell biology^20^. However, these studies were conducted exclusively in adult or experimental settings and did not capture the biology of pediatric CP, where developmental context, genetic etiology, and age-specific immune maturation may influence disease mechanisms. Critical gaps therefore remain. The immune cell composition and activation states in pediatric CP pancreatic tissue are undefined. The diversity and functional specialization of fibroblast populations, key orchestrators of pancreatic fibrosis, have not been characterized in pediatric CP. Furthermore, the transcriptional landscape of early neoplastic populations such as acinar-to-ductal metaplasia (ADM) and pancreatic intraepithelial neoplasia (PanIN) has never been examined in children, despite the markedly elevated lifetime cancer risk in this population ^21^.

To address these critical knowledge gaps, we aim to define the cellular landscape of pediatric CP using snRNA-seq, identify cell populations in diseased pancreas, characterize disease-associated transcriptional states, including fibroblast heterogeneity, immune remodeling, and presence of early neoplastic signatures. Complementing tissue-level profiling, we performed flow cytometry on peripheral blood cells to assess systemic immune alterations associated with pediatric CP. This integrative approach addresses a significant unmet need in pediatric pancreatology and provides a molecular foundation for understanding tissue-level injury paired with systemic inflammatory cell activation for future therapeutic development and long-term disease management.

## Methods

### Human pancreas samples

The study participants were enrolled after approval by the Institutional Review Board (IRB) at Cincinnati Children’s Hospital Medical Center (CCHMC) for the Total Pancreatectomy Cohort Study (IRB 2016-9510). All participants provided written informed consent to participate in this study, and tissue for research was collected at the time of Total Pancreatectomy Islet Autotransplantation (TPIAT). Pancreatic tissues were obtained from healthy pediatric human subjects (Network for Pancreatic Organ Donors; NPOD). CP was defined according to the published criteria of the International Study Group of Pediatric Pancreatitis: In Search for a CuRE (INSPPIRE). Diagnosis required characteristic imaging features of CP, including findings suggestive of CP such as, but not limited to, ductal calcifications or pancreatic ductal dilation, in addition to at least one of the following: (1) pancreatic-type pain, (2) evidence of endocrine dysfunction, or (3) evidence of exocrine dysfunction.^56^ Sample pancreatic tissues were collected for research, under minimal ischemia time, transferred into cryovials, and immediately cryofrozen in vapor phase liquid nitrogen for 10 minutes. For long-term storage, the tissues were transferred to liquid nitrogen storage tanks. In this study, pancreatic tissues from 15 CP subjects (median age: 15 years, 53.7% male) and 6 HC donors from the nPOD (median age: 14 years, 83.3% male) were used. Both groups had similar body mass index (BMI). Subject demographics are listed in **Table 1**.

**Table 1:** Subject demographics of participants for snRNAseq.

|  | **CP**<br>**n=15** | **HC**<br>**n=6** | **P-value** |
| --- | --- | --- | --- |
| **Site**<br>**CCHMC**<br>**NPOD** | 15 (100%)<br>0 (0%) | 0 (0%)<br>6 (100%) | - |
| **Age (years)** | 15 (10.0-16.0) | 14.0 (13.0-16.0) | 0.767 |
| **Sex**<br>**Male** | 8/15 (53.3%) | 5/6 (83.3%) | - |
| **BMI** | 21.5 (18.6-26.8) | 23.8 (19.4-30.0) | 0.45 |
| **Race**<br>**White/Caucasian**<br>**Black/African American**<br>**Other** | 15/15 (100%)<br>-<br>- | 4/6 (67%)<br>2/6 (33%)<br>- | - |
| **Genetics (positive)**<br>**PRSS1**<br>**CFTR**<br>**SPINK1**<br>**Others** | 5/15 (33.3%)<br>5/15 (33.3%)<br>4/15 (26.6%)<br>1/15 (6.6%) | NA | - |

Data presented as n (%) or median (25<sup>th</sup>-75<sup>th</sup> percentile).

### Nuclei isolation from human pancreas tissue

We modified a nuclei isolation protocol for human pancreas in this study following “Humphrey’s” protocol for human kidney tissue.^57,58^ Fresh nuclei lysis buffer (NLB) was prepared using 10 mL of Nuclei EZ lysis buffer (Sigma-Aldrich) with one tablet of cOmplete™ ethylenediaminetetraacetic acid (EDTA)-free ULTRA protease inhibitor cocktail (Roche). NLB-1 buffer cocktail was then prepared using 1.8 mL of NLB buffer mixed with 20 µL Protector RNase Inhibitor (Roche), 20 µL SUPERase·In™ RNase Inhibitor (Invitrogen), 20 µL of 10% BSA (in phosphate buffered saline, PBS), and 140 µL of 250 mM citric acid. Nuclei suspension buffer (NSB) was prepared using 1.8 mL PBS, 20 µL Protector RNase Inhibitor, and 180 µL 10% bovine serum albumin (BSA). Briefly, each snap-frozen tissue was placed in a 60 mm sterilized cell culture dish. Next, 1 mL of NLB-1 buffer was added, and the tissue was cut into smaller pieces (<8 mm^3^) using a sterile blade. The minced tissue was then transferred to a sterile Dounce homogenizer (Bellco), and another 1 mL of NLB-1 buffer was added. The tissue was minced 15 times with the loose pestle and then passed through a 100 µm cell strainer (Greiner Bio-One) into a 15 mL conical tube. The homogenate was re-transferred to the Dounce homogenizer and ground 7 times using the tight pestle. The homogenate was then passed through 40 µm cell strainer into a fresh 15 mL conical tube. The tube was centrifuged at 500 *g* for 5 min at 4°C. The supernatant was carefully discarded, and the nuclei were resuspended in 800 µL NSB. The suspended nuclei were passed through a 10 µm cell strainer. The isolated nuclei were observed for their quality, and the final suspension of nuclei was prepared for a count of 1000-2000 nuclei per µL, which then underwent further processing.

### Sample processing, library preparation, and sequencing

SnRNA-seq was performed at the Cincinnati Children’s Hospital Medical Center (CCHMC) Single Cell Genomics Facility (RRID:SCR_022653) according to the manufacturer’s instructions using Chromium Single Cell 3’ Reagent Kits (v3.1 or v4, Dual Index; 10x Genomics). Briefly, ∼16000-17000 nuclei were resuspended in the reverse transcription master mix and loaded with partitioning oil and gel beads into a chip to generate gel bead-in-emulsions (GEMs). Within each GEM, poly-A RNA was captured and reverse transcribed into cDNA, incorporating an Illumina TruSeq R1 primer sequence, a unique molecular identifier (UMI), and a 10x barcode. Barcoded cDNA was purified using Silane DynaBeads and amplified through 12–14 cycles of polymerase chain reaction (PCR). Full-length cDNA was enzymatically fragmented, size-selected, adapter-ligated, and further amplified to construct sequencing libraries. During library construction, P5, P7, i7, and i5 sample indexes, along with the TruSeq Read 2 primer, were added. Libraries were pooled and sequenced at Genomics Sequencing Facility (RRID: SCR_022630) at CCHMC on a NovaSeq X Plus using a 10B flow cell with the following parameters: R1: 28 cycles, i7: 10 cycles, i5: 10 cycles, R2: 90 cycles.

### Data analysis

Filtered gene-barcode expression matrices were generated from raw FASTQ files using 10X Genomics Cell Ranger (cellranger/7.0.1) with default parameters and the human reference genome hg38/GRCh38. All downstream analyses were performed using Scanpy (1.10.1).^59^ Doublets were identified and removed using the scanpy.pp.scrublet function. Low-quality cells were removed if they exhibited more than 2% mitochondrial gene expression or fewer than 100 detected genes. Next, we removed genes that were expressed in less than three cells. Additionally, cells with the number of detected genes (*n_genes_by_counts*) falling outside the 2nd and 98th percentiles were excluded from the analysis. After quality control, the remaining count matrix was log-normalized and used for dimensionality reduction and unsupervised clustering analysis. Briefly, the top 2,000 highly variable genes were selected, and gene expression was projected onto 30 principal components. Batch effects between individual samples were corrected using the Harmony algorithm.^60^ Following batch correction, the integrated data were further reduced and embedded into two-dimensional space using Uniform Manifold Approximation and Projection (UMAP), and 15 nearest neighbors were used to define the local neighborhood size. Clustering was performed using the Leiden algorithm with a resolution of 0.5, allowing separation of known cell populations based on established marker gene expression.

### Cell annotation

Cell clusters were identified based on common marker genes for various cell types in the human pancreas. General cell types: acinar cell (*PNLIPRP1, GP2, CPA1, AMY2B, AMY2A, CXCL12, RBPJL*), ductal cell (*CFTR, HNF1B, SOX9*), ADM (*SOX9, MUC1, KRT19*), PanIN (*CLDN18, MUC5AC, KRT19*), alpha cell (*GCG, ARX*), beta cell (*INS, IAPP*), delta cell (*SST*), adipocyte (*FABP4, LEP*), vascular endothelial cell (*PECAM1, VWF, IL6*), lymphatic endothelial cell (*PROX1, LYVE1*), quiescent stellate cell (*COL1A2, PDGFRA*), myofibroblast (*ACTA2, MYH11, MYL9, HOPX, POSTN, TPM1, TPM2*), inflammatory fibroblast (*PDGFRA, CXCL12, CFD, DPT, LMNA, HAS1, CXCL2, IL1R1, LIF*), Schwann cell (*SOX10, S100B*), immune cell (*PTPRC, MS4A1, CD3E, ITGAM, CD163*). Immune cells: B cell (*MS4A1, BANK1, PAX5*), memory T cell (*CCR7, LEF1, TCF7, SELL, IL7R, CD3E*), tissue-resident memory T cell (*ITGAE*), Th17 (*RORA, IL23R*), Treg (*FOXP3, IL2RA, CTLA4*), NK (*NCAM1, ZBTB16, GNLY, GZMA, PRF1*), inflammatory monocyte (*IL1B, S100A8, S100A9*), M2 macrophage (*CD163, MRC1, HMOX1, MSR1*), cDC (*ITGAX, CD74, CIITA*), pDC (*CLEC4C, IL3RA*), mast cell (*IL18R1, KIT, TPSAB1*).

### Gene set enrichment analysis

Gene set enrichment analysis (GSEA) was performed to identify pathways associated with transcriptional changes in ADM, PanIN, and fibroblast populations in CP samples. Differential gene expression for each cell type was obtained from the snRNA-seq data using the Scanpy *rank_genes_groups* function. For each cell type, genes were ranked according to their log fold changes relative to all other cells in the dataset. Ribosomal protein genes were excluded to reduce bias from highly abundant housekeeping transcripts. The remaining genes were sorted in descending order of log-fold changes to generate a ranked list for each cell type.

Pre-ranked GSEA was conducted using the GSEApy package (V1.1.11).^61^ Pathway annotations were derived from the WikiPathways gene set (c2.cp.wikipathways.v2025.1.Hs.symbols) and Reactome gene set (c2.cp.reactome.v2024.1.Hs.symbols.gmt). For each ranked gene list, enrichment scores were computed using 1,000 permutations with a fixed random seed to ensure reproducibility. Gene sets of all sizes were considered (minimum size=1, maximum size unrestricted). Statistical significance was assessed using the false discovery rate (FDR). Pathways with FDR < 0.25 were considered significantly enriched. Enrichment results were visualized using dot plots to summarize pathway significance.

### Ligand-receptor interactions

Ligand-receptor (L-R) interactions were inferred using the LIANA (v1.5.1) implementation of the CellChat method with the CellChatDB database. For comparative analysis between HC and CP, we focused on immune-to-exocrine signaling. In HC, interactions were subset to those with immune sources (B cells, T cells, myeloid cells) and exocrine cells (acinar cells, ductal cells) with CellChat p-values < 0.05. In CP, immune sources included B cells, T cells, myeloid cells, and inflammatory fibroblasts and targets included acinar cells, ductal cells, PanINs, and ADM; filtering was again performed with CellChat p-values < 0.05.

### Histology

One representative hematoxylin and eosin (H&E) slide from a formalin-fixed paraffin embedded block was obtained from each of the available CP pancreas resection samples (n=13), and whole slide imaging was performed using Leica Aperio AT2 scanning system. The slides were reviewed by an expert gastrointestinal pathologist using Aperio WebViewer, who was blinded to the snRNAseq data, and the presence of ADM, PanIN, and fibrosis were recorded. ADM was scored as a percentage of the total tissue. PanIN was graded as low or high grade and scored as absent, focal/rare foci (<5%) or frequent (>30% of duct involvement). Fibrosis was scored as a percentage of parenchymal replacements.

### Flow cytometry of human PBMCs

Peripheral blood samples were collected from eight pediatric individuals with CP undergoing TPIAT and seven age-matched HCs. Peripheral blood mononuclear cells (PBMCs) were isolated by density gradient centrifugation using Ficoll-Paque PLUS (Cytiva). The cells were washed and stored in liquid nitrogen until further use. For flow cytometry, cells were thawed, washed twice with phosphate-buffered saline (PBS), and resuspended in staining buffer (PBS + 2% fetal bovine serum). Cells were first incubated with Fc blocking reagent (BD Biosciences, Cat# 564219) for 10 minutes at 4°C to minimize non-specific antibody binding. Surface staining was performed using a comprehensive 23-color antibody panel for 30 minutes at 4°C in the dark. The panel included markers for: T cell subsets (CD3-BV510, CD4-Spark Blue 550, CD8a-PerCP, CD45RA-BUV395, CCR7-BV421, CD69-BV480), B cells (CD19-BUV563, CD20-BV785), monocytes (CD14-BV605, CD16-BUV496), dendritic cells (CD11c-BUV805, CD123-PE), NK cells (CD56-APC-R700), activation and co-stimulatory markers (CD80-AF647, HLA-DR-APC-Cy7, CCR2-BUV661), and a viability dye (Live/Dead Blue, Invitrogen) in the presence of brilliant violet stain buffer. Following surface staining, cells were fixed and permeabilized using the Fixation/Permeabilization Concentrate and diluent (eBioscience™) according to the manufacturer’s protocol. Intracellular staining was performed with antibodies against transcription factors: FOXP3-eFluor450, T-bet-AF488, RORγt-PE-CF594, and GATA3-APC in 1x permeabilization buffer for 30 minutes at 4°C. After staining, cells were washed twice and resuspended in staining buffer for acquisition. Samples were acquired using Cytek^®^ Aurora flow cytometer equipped with five lasers (355, 405, 488, 561, and 635 nm). Data analysis was performed using FlowJo software (version 10.10.0, BD Biosciences) and GraphPad Prism (version 10.6.1).

## Results

### Single nucleus transcriptomics reveal heterogeneity in cellular landscape of pediatric CP and HC pancreas

To characterize the cellular landscape and explore the cell populations, we obtained pancreas tissues from patients with CP (n = 15) and HC (n = 6) to perform snRNA-seq. A schematic representation of the workflow is shown in **Figure 1.A**. Following a standard snRNA-seq data analysis pipeline, unsupervised clustering of cells from HC samples revealed 11 distinct cell populations representing all major cellular compartments of the pancreas (**Figure 1.B**). Exocrine cells predominated with acinar (*PNLIPRP1, GP2, CPA1, AMY2A/2B, RBPJL*) and ductal (*CFTR, HNF1B, SOX9, PROX1*) cells accounting for ∼85% of cells, with alpha (*GCG, ARX*), beta (*INS, IAPP*), and delta (*SST*) endocrine cells accounting for ∼5% of total population. Stromal populations (stellate cells: *PDGFRA, COL1A2*; and myofibroblasts: *MYH11, ACTA2*) collectively accounted for approximately 1-4% of cells, whereas vascular (*PECAM1, VWF*) and lymphatic (*PROX1, LYVE1*) endothelial populations together represented approximately 1-3%, with Schwann cells (*SOX10, S100B*) comprising <1% of cells. Immune cells (*PTPRC, MS4A1, CD3E, ITGAM, CD163*) were present at low levels (∼4% combined) across individuals (**Figure 1.C**). Notably, the cellular composition was remarkably consistent across all six HC individuals, with minimal inter-individual variation, suggesting that the baseline cellular architecture of the pediatric pancreas is tightly regulated and stable across individuals in the absence of disease.

**Figure 1.**
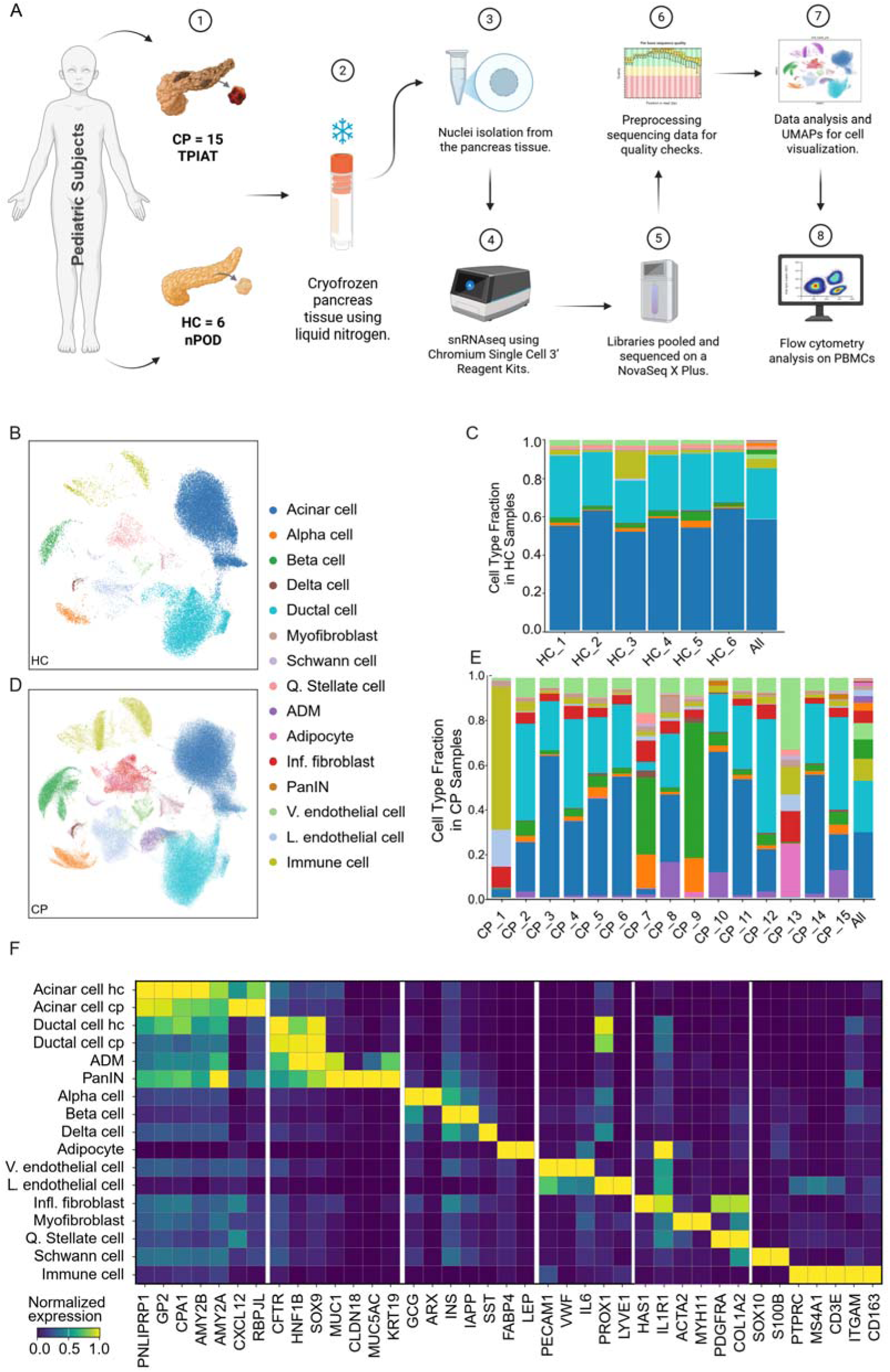
snRNA-seq analysis revealed cell type composition and marker genes expression in healthy control (HC) and chronic pancreatitis (CP) pancreatic tissues. (A) Schematic representation of the study shows tissues were obtained from 15 individuals with chronic pancreatitis (CP) undergoing total pancreatectomy with islet autotransplantation (TPIAT) and 6 healthy controls (HC) from the network of pancreatic organ donors (nPOD). Tissues were cryopreserved, and nuclei were isolated followed by snRNA-seq using the Chromium Single Cell 3’ platform (10x Genomics). Libraries were pooled and sequenced using a NovaSeq X Plus system. Following quality control and pre-processing, data were analyzed. Flow cytometry analysis of peripheral blood mononuclear cells (PBMCs) was also performed. (B, D) UMAP plots of annotated cell types in HC individuals (B) and pediatric CP individuals (D). Each color represents a distinct cell type, including acinar, endocrine (α, β, δ), ductal, endothelial (vascular and lymphatic), immune, stromal (stellate, myofibroblast, inflammatory fibroblast), Schwann cells, adipocytes, ADM, and PanIN populations. (C, E) Stacked bar plots showing the cell type fractions across HC individuals (C) and CP individuals (E). The “All” column summarizes average cell type proportions per condition. (F) Matrix plot of normalized expression levels of canonical marker genes across major cell types in HC and CP pancreatic tissues. Columns represent representative marker genes (e.g., *PNLIPRP1, GP2, CPA1* for acinar cells; *CFTR, HNF1B* for ductal cells; *INS, IAPP* for β-cells; *PECAM1, VWF* for endothelial cells; *COL1A2, PDGFRA* for stromal cells; *MS4A1, CD3E, CD163* for immune populations). Rows indicate cell type groups, and the color scale denotes normalized gene expression intensity.

In CP, the populations expanded to 15 clusters with marked heterogeneity (**Figure 1.D-E**), featuring a ∼46% reduction in acinar cells and significant remodeling of stroma and vasculature, including reduced quiescent stellate cells and enrichment of myofibroblasts and inflammatory fibroblasts (*HAS1, IL1R1*), alongside a ∼3-fold increase in vascular endothelium. Additionally, disease-associated populations emerged uniquely in CP, including ADM and PanIN, highlighting ongoing tissue injury and malignant differentiation processes towards PDAC. Expression of marker genes was non-overlapping and lineage-consistent across clusters in all identified populations (**Figure 1.F**). Cell fraction comparisons between different populations in CP and HC are depicted in **Supplementary Figure 1 and Table 1**.

### Identification of major immune cell landscape remodeling in CP

Our snRNA-seq profiling revealed major restructuring of the immune compartment in individuals with CP compared to HC. We observed that immune populations were relatively low in HC, with minimal presence of cells involved in immune responses or inflammation, such as B cell, NK, T cell, Th17, Treg, cDC and pDC. Most of the T cells were tissue-resident T cells. Macrophage compartment consisted of both M1 and M2 phenotypes. CP samples exhibited a marked increase in fraction and diversification of both lymphoid and myeloid lineages, reflecting a shift toward a highly inflammatory microenvironment in CP (**Figure 2.A**). Although cell-type annotation confirmed conserved lineage identities across conditions (**Figure 2.B**), we identified disease-specific shifts in both cell abundance and state. Notably, the CP immune microenvironment was characterized by a significant increase in inflammatory monocytes, B cells, NK cells, multiple dendritic cell subsets, along with enrichment of memory and regulatory T cell populations (**Figure 2.A, C**).

**Figure 2.**
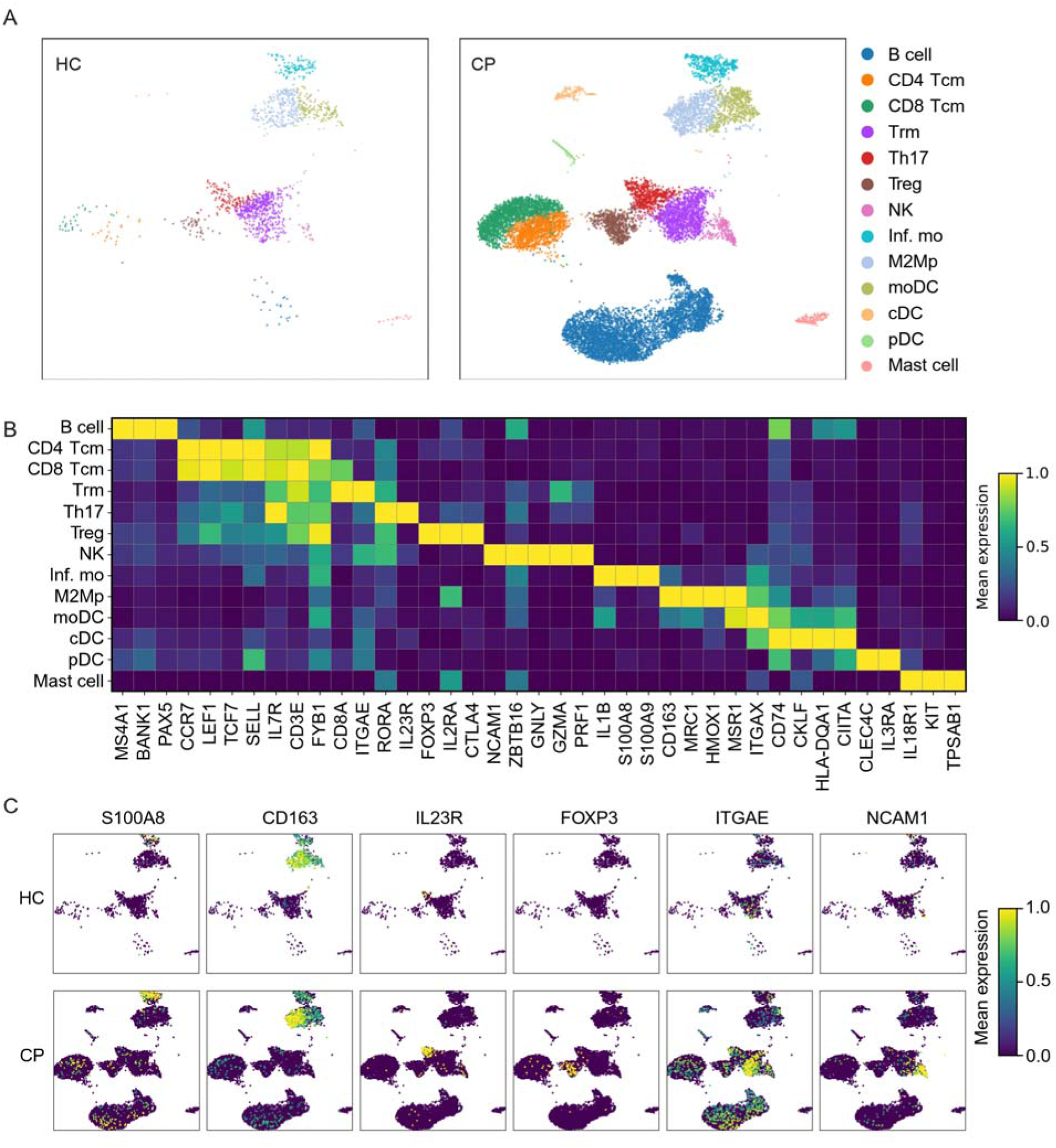
Immune cell landscape and marker gene expression in HC and CP samples. **(A)** UMAP plots of immune cell populations from HC (left) and CP (right) samples. Major immune subtypes include B cells, CD4 effector memory T cells (CD4 Tem), CD8 effector memory T cells (CD8 Tem), tissue-resident memory T cells (Trm), Th17 cells, regulatory T cells (Treg), natural killer (NK) cells, inflammatory monocytes (Inf. mo), M2 macrophages (M2Mp), monocyte-derived dendritic cells (moDC), conventional dendritic cells (cDC), plasmacytoid dendritic cells (pDC), and mast cells. **(B)** Matrix plot of normalized expression of canonical immune marker genes across immune subtypes in HC and CP samples. Markers include *MS4A1, BANK1, PAX5* (B cell markers); *CCR7, LEF1, TCF7, SELL, IL7R* (T cell subsets); *GNLY, GZMA, PRF1* (NK markers); *IL1B, S100A8, S100A9* (inflammatory monocytes); *CD163, MRC1, HMOX1* (M2 macrophages); *CD74, HLA-DQA1, CIITA* (dendritic cells); *CLEC4C, IL3RA* (pDC); and *TPSAB1* (mast cells). The color scale denotes normalized gene expression intensity. **(C)** UMAP plots showing expression patterns of representative immune markers: *S100A8, CD163, IL23R, FOXP3, ITGAE,* and *NCAM1* in HC and CP immune populations. Color scale indicates normalized mean gene expression (0–1).

This cellular reorganization was driven by fundamental alteration in functional gene expressions (**Figure 2.C**). In HC, myeloid cells were predominant with homeostatic signatures such as *CD163* and relatively low expression of inflammatory gene *S100A8*. In contrast, the CP myeloid compartment showed redistribution or differentiation into different immune subsets, with elevated expression of *S100A8/A9*, marking an increased inflammatory monocyte population (9.8%) and antigen presenting populations against HC including cDCs, and pDCs which were detectable only in CP. Mast cells were increased two-fold in CP vs. HC. Among the lymphoid populations, increased *FOXP3* marked regulatory T cells which were three-fold higher in CP vs. HC. The enhanced *CD163* could define M2-macrophage population which was mostly similar in CP and HC. Concurrently, T cell subsets in CP displayed increased *IL23R* and *FOXP3* expression, highlighting the coexistence of regulatory and Th17-like immune programs, whereas elevated *ITGAE* (CD103) and *NCAM1* (CD56) marked tissue-resident memory T cells and NK cells (two-fold increase in CP vs. HC), respectively. There was also a prominent fifteen-fold increase in CD4^+^ and CD8^+^ central memory T cell populations which were marked by increased expression of *CCR7, SELL, IL7R*. B cells were marked by *MS4A1* (CD20) and were significantly increased in CP vs. HC (fifty-four-fold). Collectively, these findings indicate that CP is associated with a coordinated shift from a relatively quiescent immune landscape in HC toward a complex, pro-inflammatory, and immunoregulatory network that likely contributes to chronic tissue damage, immune cell retention, and persistent immune activation within the pancreatic microenvironment.

### CP pancreas exhibit early PanIN and tumor (PDAC)-associated gene signatures

To determine whether damage-associated (ADM) and cancerous gene programs (PanIN) are present in our CP samples, we evaluated the expression of PDAC and tumor PanIN (malignant PanIN) gene signatures derived from two prior studies on adult healthy pancreas and pancreatic cancer.^22,23^ Our results showed that ADM and PanIN clusters in CP exhibited higher gene scores than those in normal pancreas for both exocrine cells (acinar and ductal cells), indicating that these ADM and PanIN populations display malignant-associated transcriptional programs. (**Figure 3.A, B, Supplementary Figure 2**). In contrast, the average expression levels of both gene sets were lower in exocrine cells from HC samples (**Supplementary Figure 3**). Quantification of tumor gene scores across individual CP samples revealed marked inter-patient heterogeneity in cell populations (**Figure 3.C**). ADM gene scores were >10% in three subjects (CP_8, CP_10, CP_15) while most samples showed low baseline scores (<0.25) across all cell types, indicating that tumor-associated signatures are not uniformly present but emerge in most patients, potentially reflecting differences in disease severity, duration, or progression stage. To identify biological pathways that may be associated with ADM and PDAC-associated transcriptional programs, we performed gene set enrichment analysis (GSEA) within ADM cells and PanINs respectively. ADM cells showed significant enrichment in cancer-associated and metabolic pathways (**Figure 3.E**). The top enriched pathway was pancreatic cancer subtypes (NES∼2.4, FDR<1×10^-15^), directly linking ADM to pancreatic cancer biology and suggesting that ADM represents an early step in neoplastic transformation. Additional enriched up-regulated pathways in ADMs included multiple drug metabolism and biosynthetic processes, such as glucuronidation, pregnane receptor pathway, constitutive androstane receptor pathway, codeine and morphine metabolism, estrogen metabolism, mineralocorticoid biosynthesis, bile acid synthesis, eicosanoid metabolism, and irinotecan pathway, which suggested aberrant metabolic activities in ADM cells.

**Figure 3.**
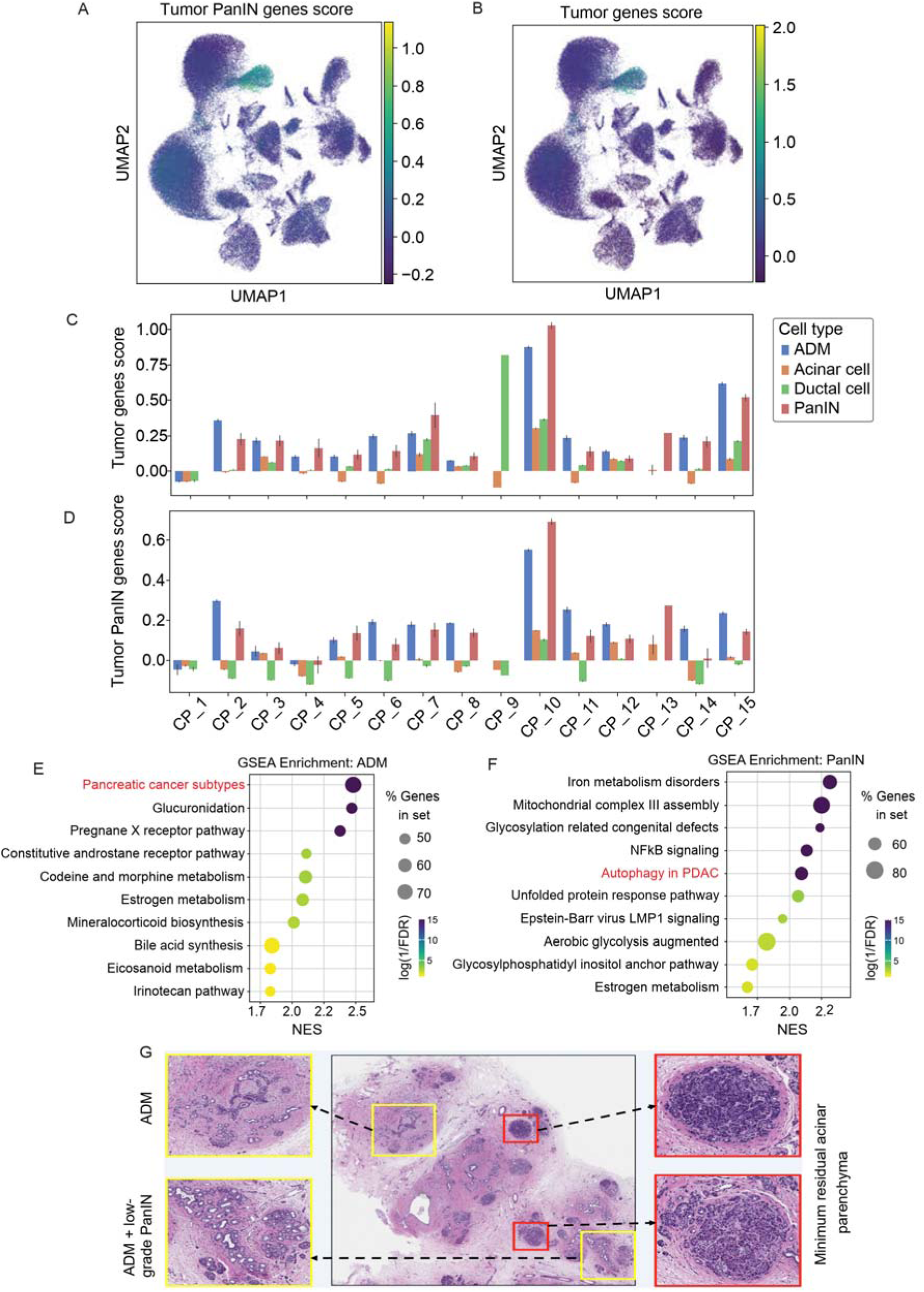
PanIN and tumor-related gene scores across CP patients. **(A-B)** UMAP projection of pancreatic cells’ colors by tumor PanIN gene score **(A)** and tumor (PDAC) gene score **(B)**, showing tumor PanIN and PDAC signature enrichment across PanIN and ADM populations in CP. **(C)** Tumor gene scores across exocrine cell types in each individual CP sample, highlighting elevated tumor gene signatures in ADM and PanIN populations relative to normal acinar and ductal cells. **(D)** Tumor PanIN gene scores across exocrine cell types in each individual CP sample, revealing selective enrichment of PanIN-associated gene expression within specific epithelial subsets. **(E–F)** Gene set enrichment analysis (GSEA) of consensus programs enriched in ADM **(E)** and PanIN **(F)** populations. Dot size indicates the percentage of genes in each pathway; color represents −log10(FDR), and the x-axis shows normalized enrichment score (NES). Enriched pathways reflect pancreatic cancer–related signaling in ADM and PanIN. **(G)** Histology of CP pancreas tissue revealed atrophic acinar cells with minimal residual acinar parenchyma as shown in the red boxes. There was reduced acinar cell density compared to normal pancreatic tissue. There was extensive interstitial fibrosis with inflammatory infiltrates and significant loss of normal pancreatic parenchyma. The presence of ADM was clearly observed with inclusion of low-grade PanINs as shown by the yellow boxes on the left.

Analysis of tumor PanIN gene scores across all 15 CP individuals revealed a similar variability, with two individuals (CP_10, CP_15) exhibiting high tumor PanIN gene scores (**Figure 3.D**). GSEA of PanIN cells identified enrichment of pathways associated with cellular stress, altered metabolism, and cancer progression (**Figure 3.F**). The most interesting statistically significant pathway was autophagy in pancreatic adenocarcinoma (NES∼2.15, FDR<1×10^-10^), suggesting that autophagy-related transcriptional programs may be active in PanIN populations within CP. These findings are consistent with prior reports implicating autophagy in pancreatic tumor biology; however, functional validation is required to determine its precise role in pediatric CP-associated PanIN lesions.^24,25^ Additional enriched pathways included iron metabolism disorders (NES∼2.35), mitochondrial complex-III assembly, glycosylation related congenital defects, NFκB signaling, photodynamic therapy- induced unfolded protein response, Epstein-Barr virus LMP1 signaling, aerobic glycolysis augmented, glycosylphosphatidyl inositol anchor pathway, and estrogen metabolism.

To validate our transcriptomic analysis in terms of pathological aspects, we analyzed H&E-stained CP pancreatic tissue sections from 13 cases (**Figure 3.G**). Histological examination confirmed the presence of ADM and/or low-grade PanIN lesions in 9 of the CP tissue samples (69%) (**Supplementary Table 2**). ADM was defined as foci of atrophic acinar parenchyma with a phenotypic transition to small, ductal-like morphology, lacking significant cytologic atypia or mucin production. Low-grade PanIN lesions were identified by their characteristic ductal flat epithelial proliferations containing mucin producing cells with basally located, small oval nuclear and minimal cytologic atypia. No high-grade PanIN were identified in any of the tissue sections. In most cases (n=5), ADM was relatively patchy (30% or less total tissue area); however, two cases showed 50% and >75% replacement. PanIN was very focal in all cases except for one, in which there was near-total replacement of parenchyma by fibrosis with frequent foci of low-grade PanIN. Interestingly, the two cases with high ADM and PanIN expression from the snRNAseq analysis (CP_10, CP_15) showed none to minimal ADM or PanIN on histological assessment. This assessment might reflect that molecular features may precede histologically identifiable alterations or this might be due to a sampling bias, since the tissue for snRNAseq analysis was not the same segment as the one assessed histologically.

### Emergence of inflammatory fibroblast population in CP

In the normal HC pancreas, myofibroblasts and stellate cells comprised 24.7% and 75.3% of the total fibroblast fraction, respectively. In CP, the total fibroblast population increased by 57% compared to that in HC, resulting in altered ratios among fibroblast subtypes. Specifically, the stellate cell population decreased in CP to 8.3%, while myofibroblasts remained consistent at 24.7% compared to HC, and inflammatory fibroblasts emerged to 69.2%, as compared to HC. Each fibroblast population exhibited distinct marker gene expression. Myofibroblasts expressed *ACTA2, MYL9, HOPX, POSTN, TPM1*, and *TPM2* at high levels. Stellate cells were characterized by expression of *THBS1,* elevated *COL1A2*, and *CFD*. Inflammatory fibroblasts demonstrated higher expression of chemokines, including *CXCL2, CXCL12*, and, genes previously associated with inflammatory fibroblasts, such as *PDGFRA, IL1R1, LIF*, *HAS1*, *LMNA*, and *CFD* **(Figure 4.A)**. Histology of tissues confirmed the presence of fibrosis in 85% of samples with at least five tissue slides showing >30% fibrosis score (**Supplementary Table 2**). GSEA analysis suggested that both myofibroblasts and inflammatory fibroblasts are involved in extracellular matrix (ECM) remodeling during CP **(Figure 4.B, Supplementary Figure 4.A, B)**.

**Figure 4.**
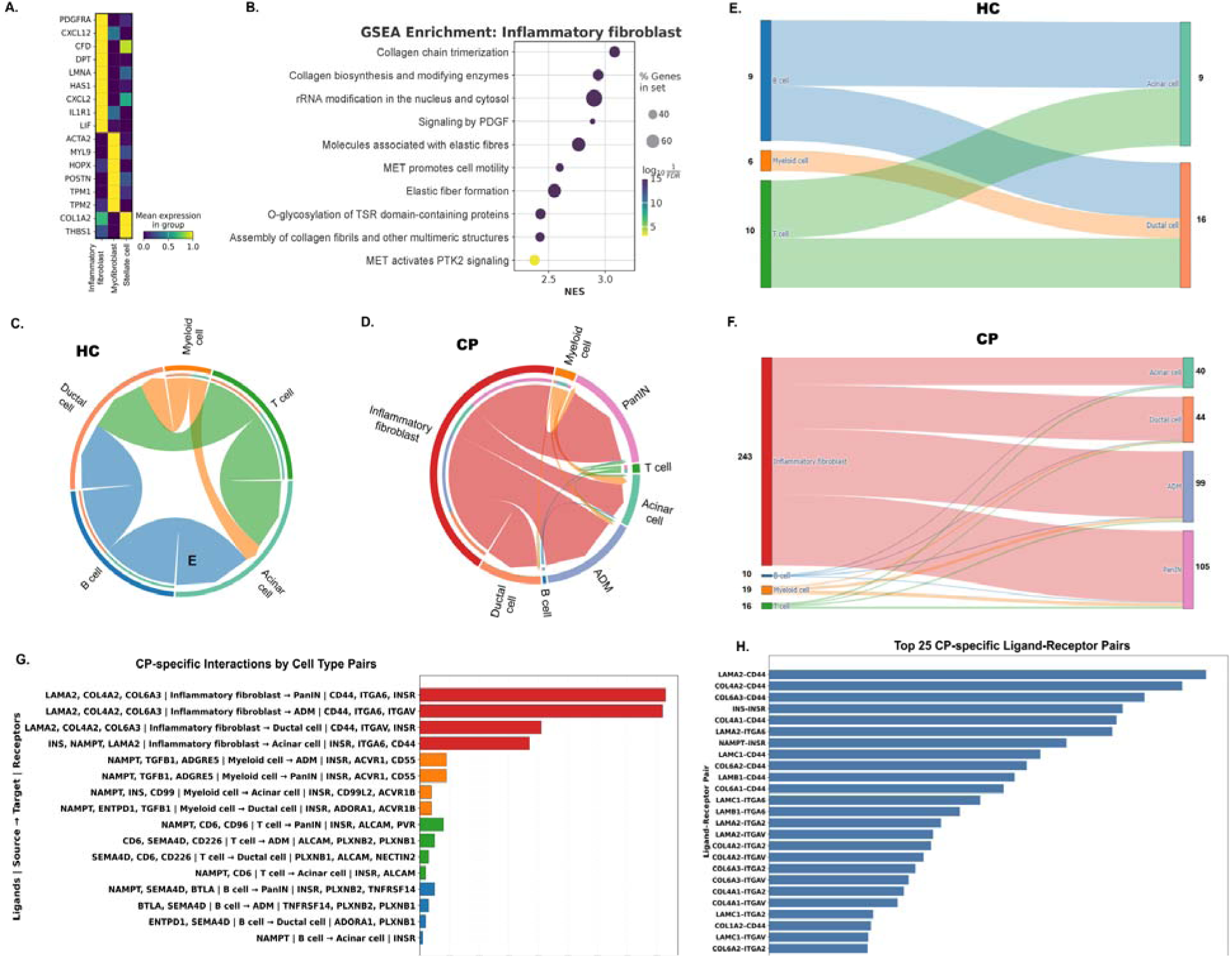
Fibrosis and cell-cell interactions. **(A)** Matrix plot showing mean expression of marker genes across three fibroblast subpopulations identified in CP: inflammatory fibroblasts, myofibroblasts, and stellate cells. Inflammatory fibroblasts were characterized by high expression of nine markers; myofibroblasts by six markers; and stellate cells by three markers. Color scale represents normalized mean gene expression (0-1). **(B)** Dot plot showing Gene Set Enrichment Analysis (GSEA) of pathways enriched in inflammatory fibroblasts from CP tissue. Dot size indicates the percentage of genes in each pathway; color represents −log10(FDR), and the x-axis shows normalized enrichment score (NES). Top enriched pathways included collagen chain trimerization, collagen biosynthesis, rRNA modification, PDGF signaling, and elastic fiber formation. **(C)** Chord diagram showing cell-cell communication networks in HC based on CellChat analysis of ligand-receptor (L-R) interactions. Colored outer arcs represent cell types (Myeloid, T, Acinar, B, Ductal cells); inner ribbons indicate signaling interactions between cell populations, with ribbon width proportional to interaction strength. The dominant interaction was between Acinar cells and B cells. **(D)** Chord diagram showing cell-cell communication networks in CP showing increased number of interactions with eight distinct populations (Myeloid, T, Acinar, B, and Ductal cells including inflammatory fibroblasts, PanIN, and ADM). Inflammatory fibroblast interactions are dominant with extensive bidirectional communication to ductal cells, myeloid cells, and other cell types. **(E)** Sankey diagram showing HC-specific L-R interactions. **(F)** Sankey diagram showing CP-specific L-R interactions. Flow width indicates interaction strength. **(G)** CP-specific L-R interactions ranked by number of interactions. The interactions were grouped by source cell type (inflammatory fibroblasts, myeloid, T, and B cells). Inflammatory fibroblasts showed the strongest CP-specific signaling to PanIN, ADM, and ductal cells, primarily through laminin/collagen-integrin interactions (*LAMA2*, *COL4A2*, *COL6A3, INS, NAMPT* → *CD44*, *ITGA6*, *INSR*, *ITGAV*). **(H)** Top 25 CP-specific L-R pairs by interaction probability across all cell type pair combinations. *LAMA2* → *CD44* showed the highest probability (0.1), followed by *COL4A2* → CD44, *COL6A3* → *CD44, and INS* → *INSR*.

### Cell-cell interactions reveal higher ligand-receptor pairs between immune and exocrine cells in CP

To characterize intercellular signaling alterations during pancreatic inflammation, we performed CellChat analysis on immune and exocrine cell populations from HC and CP tissue samples. In healthy pancreas, immune-to-exocrine communication was balanced across immune compartments, B cells, myeloid cells, and T cells, with comparable mean interaction probabilities (0.14, 0.03, and 0.11, respectively) for both exocrine targets, acinar and ductal cells. Overall, we detected 35 significant interactions (p<0.05) in the HC condition (**Figure 4.C**). These interactions primarily targeted acinar and ductal cells through 25 unique L-R pairs in HC, with *PRSS1/2-PARD3* signaling representing the dominant pathway (interaction probability 0.27-0.30). Within the unique interactions for HC, T cells showed 10 interactions with *PRSS1/2*→*PARD3* as the most significant L-R pairs, and myeloid cells showed 6 interactions with *PRSS2*→*PARD3*, *INS/NAMPT*→*INSR* as the most significant L-R pairs (**Figure 4.E**). In contrast, CP tissues exhibited a marked 8.5-fold expansion of immune-exocrine signaling (298 interactions and 288 unique L-R pairs) (**Figure 4.D**). Inflammatory fibroblasts emerged as the dominant signaling source, accounting for 82% of total interactions and disease-associated ADM and PanIN appeared as targets of signaling, compared to the interaction sources and targets in HC. Within the unique interactions specific to CP, the inflammatory fibroblasts signaling preferentially targeted disease-associated cell types, with the highest interactions (243) and probabilities directed toward PanIN cells (0.019, 83 L-R pairs) and ADM (0.018, 82 L-R pairs), compared to normal ductal (0.024, 41 pairs) and acinar cells (0.017, 37 pairs) (**Figure 4.F**).

The most prominent CP-specific interactions were from inflammatory fibroblasts as source, which contributed to the highest numbers of CP-specific interactions with PanIN and ADM, followed by ductal and acinar cells. The interactions were dominated by basement membrane/adhesion axes (*LAMA2/COL4A2/COL6A3*→*CD44/ITGA6/ITGAV*) and growth factor/metabolic signaling (*INS/NAMPT*→*INSR*). In contrast, immune-derived CP-specific interactions were fewer and more target-restricted: myeloid cells mainly interacted with ADM and PanIN (*NAMPT/INS/CD99/TGFB1*→*INSR/ACVR1B/ACVR1/ACVR1B*), with smaller contributions to acinar and ductal cells; T cells showed modest interactions with PanIN and ADM with limited ductal interactions (*SEMA4D/CD226/CD6*→*PLXNB1/PLXNB2/NECTIN2/ALCAM*) (**Figure 4.G**).

Ranking CP specific L-R pairs by their interaction probability revealed a highly skewed distribution dominated by ECM-associated signaling (**Figure 4.H**). The *LAMA2*→*CD44* L-R pair emerged as the top CP specific interaction, indicating strong metabolic signaling uniquely enriched in CP relative to HC, centered on matrix engagement. This was followed by a broad cluster of basement membrane and collagen ligands including *COL4A2, COL6A3, COL4A1*, and *COL6A2*-predominantly interacting with *CD44* and integrins (*ITGA6, ITGA4, ITGA7, ITGA2*), and by metabolic/growth factor signaling (*INS*→*INSR*, *NAMPT*→*INSR*). Multiple laminin-integrin and collagen-*CD44* pairs ranked among the highest-ranked interactions, highlighting extensive ECM-receptor communication as a defining feature of CP specific signaling. Notably, 204 of 298 CP interactions (68%) involved ADM or PanIN as targets, highlighting the preferential communication between inflammatory and precancerous cells.

### Complementary flow cytometry analysis of human peripheral blood cells reveals systemic immune dysregulation

We performed a comprehensive flow cytometry analysis of peripheral blood mononuclear cells (PBMCs) from 8 pediatric CP individuals and 7 HC to corroborate and extend the tissue-level observations from snRNA-seq. Using a 23-color flow cytometry panel consisting of markers of major immune cell lineages, activation status, and co-stimulatory molecules, we quantified immune cell populations and assessed their activation states through measurements of key surface and intracellular markers using flow cytometric analysis (**Figure 5.A**).

**Figure 5.**
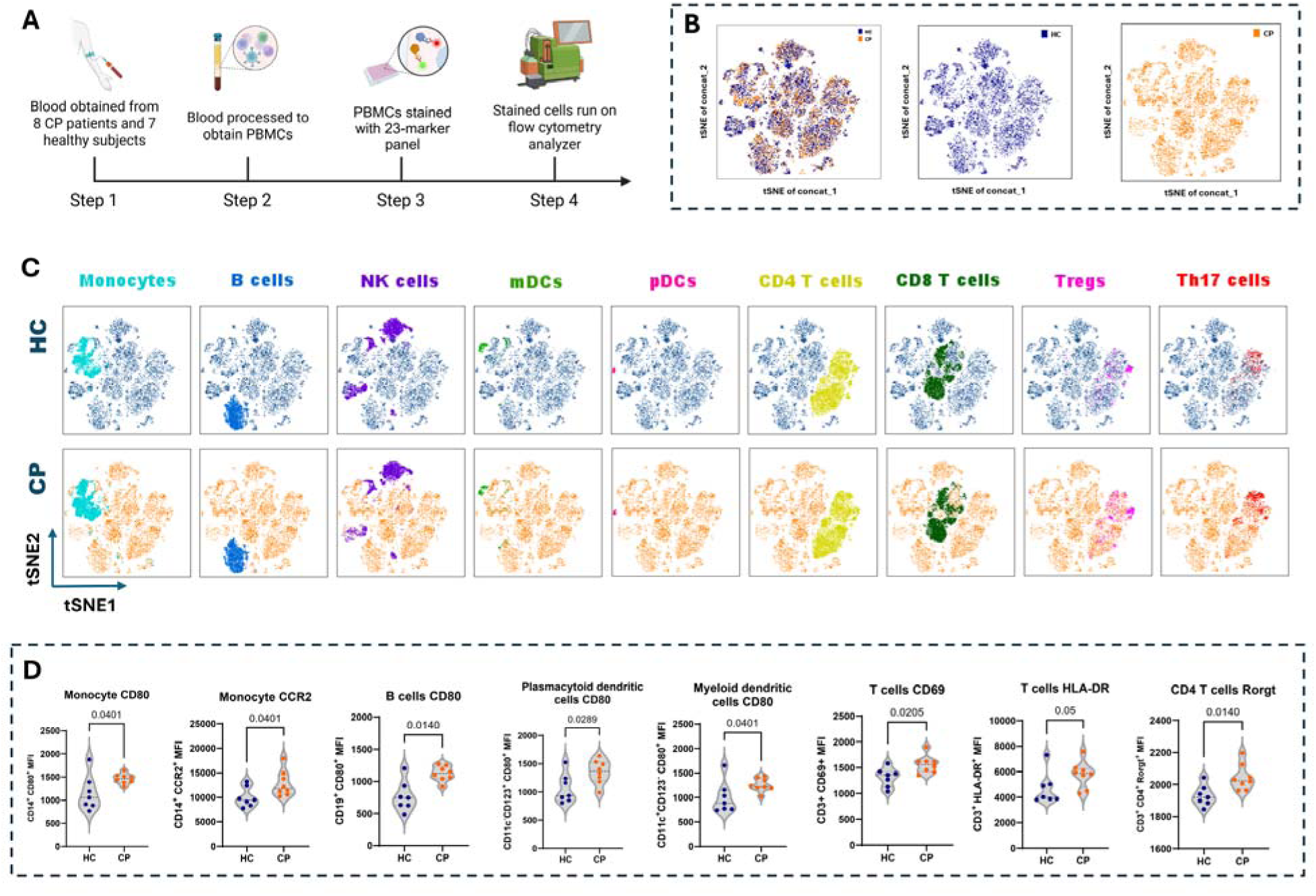
Peripheral blood mononuclear cells immune profiling using flow cytometry. **(A)** Experimental workflow for peripheral blood mononuclear cell (PBMC) analysis using flow cytometry. Blood was collected from eight CP individuals and seven HC, processed to isolate PBMCs, stained with a 23-marker flow cytometry panel, and analyzed by flow cytometry. **(B)** t-SNE visualization of concatenated samples showing combined immune cell distribution (left), HC samples (middle), and CP samples (right). Each dot represents a single cell. **(C)** t-SNE plots showing the spatial distribution of nine major immune cell subsets identified by manual gating; Monocytes, B cells, NK cells, myeloid dendritic cells (mDCs), plasmacytoid dendritic cells (pDCs), CD4 T cells, CD8 T cells, regulatory T cells (Tregs), and Th17 cells. Top row shows HC samples; the bottom row shows CP samples. **(D)** Quantitative comparison of activation marker expression between HC and CP groups. Violin plots show mean fluorescence intensity (MFI) for different immune parameters: Monocyte CD80, Monocyte CCR2, B cell CD80, pDC CD80, mDC CD80, T cell CD69, T cell HLA-DR, and CD4 T cell RORγt. Each dot represents an individual subject (HC, n=7; CP, n=8). Data are presented as mean ± standard deviation. Statistical significance was determined by Mann-Whitney U test. p-values ≤0.05 were considered statistically significant.

As shown in **Figure 5.B**, PBMCs from CP and HC showed overall similar cell type distributions, with the exception of monocytes and activated T cell populations. We identified nine major immune populations through manual gating: monocytes, B cells, NK cells, mDCs, pDCs, CD4 T cells, CD8 T cells, Tregs, and Th17 cells (**Figure 5.C**).

Significant differences were observed on performing quantitative analysis of activation markers across different immune cell populations (**Figure 5.D**). Classical monocytes (CD14^+^ cells) from individuals with CP demonstrated significantly elevated levels of the co-stimulatory molecule CD80 compared to HC (MFI: CP, 1461 ± 105 vs. HC, 1155 ± 380; p = 0.0401). Additionally, monocytes from individuals with CP exhibited significantly higher levels of the chemokine receptor CCR2 (MFI: CP, 12793 ± 2632 vs. HC, 10010 ± 2033; p = 0.0401), the primary receptor for MCP-1/CCL2 that mediates monocyte recruitment to inflamed tissues. The elevated CCR2 on circulatory monocytes aligns with the tissue-enriched inflammatory monocyte and myeloid cell expansion observed in CP tissue. B cells (CD19^+^) from CP individuals exhibited significantly increased CD80 expression compared to HC (MFI: CP, 1124 ± 116 vs. HC 768 ± 236; p = 0.014). Plasmacytoid dendritic cells (pDCs; CD123^+^CD11c^−^), which specialize in type I interferon production, from CP individuals s, showed significantly elevated CD80 levels in individuals (MFI: CP, 1354 ± 208 vs. HC, 1061 ± 260; p = 0.0289). Similarly, myeloid dendritic cells (mDCs; CD11c^+^CD123^−^), which are highly efficient at antigen presentation, demonstrated significantly increased CD80 levels (MFI: CP, 1259 ± 136 vs. HC, 1001 ± 327; p = 0.0401).

Analysis of T cell subsets revealed significant alterations in activation marker expression. CD8^+^ T cells from CP individuals displayed significantly increased expression of the early activation marker CD69 (MFI: CP, 1782 ± 335 vs. HC, 1413 ± 201; p = 0.0289), indicating recent TCR engagement. Furthermore, T cells (CD3^+^) demonstrated elevated HLA-DR expression (MFI: CP, 5778 ± 1041 vs. HC, 4681 ± 1268; p = 0.05), a marker of sustained T cell activation typically associated with chronic inflammatory and autoimmune conditions. Notably, CD4^+^ T cells from CP individuals showed elevated levels of RORγt (MFI: CP, 2044 ± 80 vs. HC 1930 ± 65; p = 0.014), the master transcription factor for Th17 cell differentiation. Both CD8^+^ central memory (TCM) and effector memory (Tem) cell populations showed evidence of activation in CP individuals. The TCM and Tem cells exhibited higher levels of CD69 in the CP group compared to HC (MFI: CP, 1432 ± 165 vs. HC, 1151 ± 236; p = 0.021 and MFI: CP, 3398 ± 1199 vs. HC, 1986 ± 428; p = 0.004 respectively). The elevated RORγt in circulating CD4^+^ T cells and increased CD69 on CD8^+^ T cells in peripheral blood mirrors the Th17 and tissue-resident T cell enrichment identified by snRNA-seq, indicating that adaptive immune dysregulation in pediatric CP extends beyond the pancreas.

## Discussion

This study presents a comprehensive single-nucleus transcriptomic atlas of pediatric CP, revealing widespread cellular heterogeneity and early neoplastic transformation that distinguish this condition from healthy pediatric pancreas. Through integrated analysis of approximately 197, 000 nuclei from pediatric CP and HC tissues, combined with peripheral blood immune profiling, we have uncovered important insights into the disease, linking local tissue inflammation with systemic immune dysregulation. Our findings suggest that pediatric CP is a highly complex disorder characterized by (1) emergence of inflammatory fibroblast populations that may drive fibrogenesis and increased cell-cell communication, (2) ADM signatures representing tissue injury and PanIN signatures representing early neoplastic transformation, (3) increased alterations in immune cell composition and activation states, and (4) dysregulated L-R interactions that may lead to chronic inflammation. These analyses may provide a molecular foundation for understanding long-term cancer risk post-pediatric CP and identifying novel therapeutic targets.

Our snRNA-seq analysis identified 18 distinct cell populations in the pediatric pancreas (both healthy and diseased), including acinar cells, ductal cells, endocrine cells (alpha, beta, delta), stellate cells, fibroblasts (inflammatory fibroblasts, myofibroblasts), endothelial cells (vascular, lymphatic), immune cells (T cells, B cells, myeloid cells, mast cells), Schwann cells, adipocytes, ADM, and PanIN. This cellular diversity aligns with recent single-cell atlases of human pancreas but reveals immune landscapes in detail that have not been previously characterized.^20,26^ We observed dramatic shifts in cellular composition in CP compared to HCs, with significant expansion of fibroblast populations and immune cells, accompanied by a reduction in acinar cells. These findings parallel observations in adult human diseased pancreas with different fibroblast and immune cell populations.^27,28^

Immune cell infiltration is a hallmark of pancreatitis; however, the specific immune populations and their functional states in pediatric CP have not been previously characterized. Our snRNA-seq analysis revealed a 2-fold increase in immune cell abundance in CP tissue compared to HC, with significant expansion of inflammatory monocytes, mast cells, central memory T cells, B cells, Th17, Treg, and NK populations, which were largely absent in HC. Specifically, we observed high number of immune cells in CP_1, which is interesting and shows that there is varied immune cell infiltration in different CP patients. This immune infiltration was accompanied by significant transcriptional changes indicating activation (*CCR7, SELL, IL18R1, IL7R, IL2RA*), differentiation (*MS4A1, FOXP3, ITGAE, IL3RA, CD163*), and effector (*GNLY, GZMA, IL1B, S100A8/9, MRC1*) function acquisition. T cells were the major immune population in CP tissue, comprising both CD4^+^ and CD8^+^ subsets. CD4^+^ T cells exhibited a mixed phenotype with expression of Th17 markers (*IL23R, RORA*), suggesting pro-inflammatory polarization. Th17 cells are critical mediators of autoimmune and chronic inflammatory diseases, secreting *IL-17A, IL-17F*, and *IL-22* to recruit neutrophils and promote tissue damage.^29^ In autoimmune pancreatitis, Th17 cells drive disease pathogenesis through IL-17-mediated activation of pancreatic stellate cells and fibroblasts.^30^ Our finding of elevated Th17 markers in CP tissue suggests that similar mechanisms operate in pediatric CP. In experimental CP, anti IL 17A treatment reduced CP severity in the absence of stimulator of interferon genes (STING), STING deficiency promoted Th17 polarization, pancreatic stellate cells expressed functional IL 17 receptors with upregulation of fibrosis genes, and CP patient pancreata contained more IL17A cells than tumor margins.^31^ Similar immune polarization has been reported in adult CP, where single cell profiling uncovered enriched CCR6 CD4 T cells, and monocyte-T cell crosstalk (CCR6-CCL20) which was previously related to pro-inflammatory cytokines and tumor-promoting role in pancreatic cancer.^32,33^ These studies are consistent with the presence of Th17 populations in our CP tissue samples. Serum proteomics from a large CP cohort singled out IL 17 axis markers as discriminators of CP along the pancreatitis continuum, consistent with our increased *IL23R/RORA/IL17R* expression in tissue Th17 cells.^34^ The prominence of *S100A8/A9* expression in inflammatory monocytes in CP reflects known roles of *S100A8/A9* in leading to leukocyte recruitment and inflammatory role, reinforcing their possibility as upstream drivers of pancreatic inflammation.^35^ Finally, the increase in *ITGAE* (CD103) expression in Trm in our study coincides with prior human pancreas data showing CD103^+^ Trm cells mediate immune homeostasis in individuals with CP, where there was an increase in T-bet levels in pancreatic CD8^+^ Trms compared to the control group, and this rise in T-bet was strongly linked to a reduction in PD-1 expression.^36^

A key strength of this study is the convergence between peripheral immune profiling and tissue transcriptomics. The systemic upregulation of monocyte activation markers, particularly CD80 and CCR2, in peripheral blood is consistent with the tissue-level enrichment of inflammatory monocytes and fibroblast-driven CCL2 signaling identified by snRNA-seq, suggesting that circulating monocytes may be actively recruited to the inflamed pancreas via the CCL2-CCR2 axis. The widespread activation of T cell compartments in our pediatric CP cohort evidenced by elevated CD69 expression across CD4^+^, CD8^+^, and memory T cell subsets indicate sustained immune stimulation. CD69 is an early activation marker rapidly upregulated upon TCR engagement.^37^ The elevated RORγt expression in FOXP3^+^ regulatory T cells from CP individuals represents important implications for immune regulation. RORγt is the master transcription factor for Th17 cell differentiation, and its expression in Tregs suggests plasticity toward a Th17-like phenotype.^38^ The Treg-to-Th17 shift may be driven by inflammatory cytokines in CP, particularly IL-6 and TGF-β, which promote RORγt expression even in FOXP3^+^ cells.^39^ Therefore, RORγt^+^ Treg population could serve as a biomarker for Th17 skewing and potentially predict individuals at higher risk for progressive disease. Together, these findings suggest a model in which chemokine gradients recruit activated monocytes and T cells from circulation into pancreatic tissue, where they encounter locally produced chemokines and cytokines that further activate and retain them. The activated DCs we identified in peripheral blood may traffic to pancreatic lymph nodes where they present pancreatic antigens to naïve T cells, generating effector and memory T cell responses that then home to the pancreas.^40^ This pathway predicts that blocking chemokine receptors (CCR2, CCR6) or co-stimulatory molecules (CD80-CD28 interaction) could interrupt the trafficking and activation cycles that perpetuate CP. Our findings in pediatric CP reveal important differences from published adult CP studies suggesting age-dependent or disease-dependent immune profiles. While adult CP is characterized by peripheral lymphopenia with age-dependent reductions in CD3^+^, CD4^+^, and CD8^+^ T cells ranging from 24% to 67%, our pediatric cohort demonstrated preserved T cell numbers among other immune cells with elevated activation markers.^41^ This suggests that pediatric CP may represent an earlier, more inflammatory stage characterized by immune activation, whereas adult CP may reflect a later stage with immune exhaustion and lymphocyte depletion. The genetic predominance in pediatric CP, with *PRSS1*, *CFTR*, *SPINK1*, and other mutations in 60-70% of cases, may also contribute to distinct immune profiles compared with adult CP.^10^ This multi-compartment immune dysregulation present both in tissue and blood, underscores the systemic nature of pediatric CP and highlights circulating immune markers as potential biomarkers for disease monitoring and therapeutic targeting.

One of the most clinically significant findings of our study is the identification of ADM and PanIN signatures in pediatric CP tissue. ADM is a cellular reprogramming process in which differentiated acinar cells lose their secretory phenotype and acquire ductal characteristics, often in response to injury or inflammation.^42,43^ PanINs are well-established precursor lesions for PDAC, classified into low-grade (PanIN-1, PanIN-2) and high-grade (PanIN-3) based on architectural and cytological atypia.^44^ The presence of both ADM and early PanIN signatures in our pediatric CP samples indicates that tissue injury or neoplastic transformation may begin early in the disease course, potentially decades before clinical cancer diagnosis. Our transcriptomic analysis revealed that ADM cells exhibit a hybrid acinar-ductal phenotype, with lower expression of acinar markers (*PRSS1, CPA1, AMY2A*) and increased expression of ductal markers (*KRT19, SOX9, MUC1*). This is consistent with previous studies showing that ADM represents a transitional state with plasticity toward either regeneration or neoplastic progression.^42,45^ PanIN signatures identified in our study exhibited more advanced neoplastic features, including expression of oncogenic markers such as *MUC5AC, CLDN18,* and *KRT19*. *MUC5AC* is a mucin glycoprotein that is normally absent in the pancreas but is aberrantly expressed in PanIN and PDAC, where it promotes tumor growth and metastasis.^46,47^ The co-expression of ADM and/or PanIN (low-grade for histology) was observed in 86% and 69% of the transcriptomic and histologically assessed CP tissues, respectively. The co-expression of these markers in pediatric CP-associated PanINs indicates that these lesions may contain pro-tumorigenic properties and represent a critical window for cancer prevention. The molecular pathways enriched in ADM and PanIN cells provide further insights into neoplastic transformation mechanisms. The enrichment of pancreatic cancer subtypes, activation of autophagy, and NF κB pathways are highly consistent with mechanistic studies showing that elevated autophagy and context dependent NF κB activity promote PanIN persistence and progression of cancer.^48,49^ Current surveillance guidelines for hereditary pancreatitis recommend initiating pancreatic cancer screening at 40-50 years of age; however, our findings suggest that neoplastic changes may begin much earlier.^50^ These findings raise the question of whether earlier surveillance or chemoprevention strategies should be considered for high-risk pediatric pancreatitis patients.

Another prominent finding of our study was the identification of inflammatory fibroblasts as the dominant fibroblast population. These cells exhibited high expression of ECM genes including *COL1A2* along with inflammatory mediators such as *CXCL2, IL1R1,* and *CXCL2*. The dual ECM-inflammatory signature distinguishes inflammatory fibroblasts from quiescent pancreatic stellate cells and positions them as central orchestrators of fibrogenesis in pediatric CP. Recent studies in adult pancreatic cancer have identified functionally distinct fibroblast subsets with divergent roles in tissue remodeling and tumor progression.^51,52^ In PDAC, cancer-associated fibroblasts (CAFs) are classified into myofibroblastic CAFs (myCAFs) characterized by high α-SMA expression and iCAFs marked by *IL6* and *CXCL1* secretion.^53^ The study also showed that IL-1 signaling was the main pathway responsible for induction of inflammatory phenotypes in CAFs. Our inflammatory fibroblasts share transcriptional features with CAFs, suggesting that chronic inflammation in pediatric CP may establish a pro-tumorigenic microenvironment years before malignant transformation. This is particularly concerning given that hereditary pancreatitis confers up to 53-fold increased PDAC risk and that our cohort includes individuals with genetic predisposition.^12^

In the field of pancreatic diseases, it is now widely recognized that cell-cell communication within the tissue microenvironment may contribute to pancreatic diseases or cancer initiation and progression.^54^ Our cell-cell communication analysis using CellChat revealed profound rewiring of intercellular signaling networks in CP compared to healthy pancreas. In healthy pancreas, cell-cell interactions were dominated by interactions to exocrine targets acinar and ductal cells. In contrast, CP was characterized by a significant expansion of immune-to-exocrine and fibroblast interactions, dominated by inflammatory fibroblasts. CP-specific cell interactions were dominated by metabolic and ECM-mediated signaling, with strong laminin (*LAMA2/LAMB1*) and collagen (*COL4A1/2, COL6A1/3*) signals to *CD44* and α integrins (*ITGA6/ITGA5/ITGAV*) showing ECM–receptor crosstalk as a defining feature of diseased pancreatic stroma. Cell communication also revealed metabolic/growth factor signaling (*INS*→*INSR* and *NAMPT* interactions), pointing to niche level metabolic adaptation consistent with evidence that autophagy and stress metabolism support epithelial survival during early pancreatic neoplasia.^24,55^

This study has several notable strengths. It represents a comprehensive single-nucleus transcriptomic atlas of pediatric CP, providing high molecular resolution of cellular composition and gene expression in this understudied population. The sample size (15 CP patients and 6 HC) is substantial for single-cell studies and provides sufficient power for identifying major cell populations and disease-associated transcriptional changes. The integration of tissue-level snRNA-seq with peripheral blood flow cytometry demonstrates systemic immune dysregulation. The identification of preneoplastic signatures (PanIN and ADM) at the molecular level provides direct evidence of early neoplastic transformation in pediatric CP and establishes a foundation for future cancer prevention studies. Cell-cell communication analysis using CellChat reveals disease-specific signaling networks and identifies multiple druggable targets, including inflammatory fibroblast-derived pathways. This study has several limitations. Potential sampling bias may affect the representativeness of the data, and the lack of longitudinal samples prevents assessment of disease progression. An interesting observation was that the molecular data on these CP tissues was not reflected in H&E slides of the tissues from the same patients, which means that preneoplastic programs can precede and exceed the visible histologic burden. It might be noted that the tissue sent for RNA seq analysis was not the same segment of tissue that was formalin fixed and reviewed by histology; therefore, some of the discrepancies seen could be explained by tissue heterogeneity as a sampling bias, which is often a limitation when dealing with representative sampled material and histologic analysis. The identified inflammatory fibroblast signatures, ADM/PanIN lesions, and proposed therapeutic targets (such as laminin/collagen-integrin pathways) may be potential drivers of chronic inflammation and injury and require further functional studies both *in vitro* and *in vivo*. Peripheral blood immune profiling may not fully reflect the pancreatic tissue microenvironment, and paired tissue-blood samples would strengthen correlations. The ADM and PanIN signatures were generally identified transcriptionally in samples but were not detected in most histological samples, and KRAS mutation analysis is needed to definitively assess neoplastic potential. Finally, findings may be specific to genetic/hereditary CP etiology and a predominantly Caucasian cohort, limiting applicability to other CP etiologies and ethnic populations.

Despite these limitations, this study provides a novel comprehensive single-nucleus transcriptomic atlas of pediatric CP, revealing important insights into fibroblast-driven pathology, early neoplastic changes, and immune dysregulation. We identified inflammatory fibroblasts as central orchestrators of fibrosis and cell-cell communication, characterized ADM and PanIN, and demonstrated concordant immune activation in pancreatic tissue and peripheral blood. Our findings establish pediatric CP as a complex disorder characterized by substantial inter-patient heterogeneity, involving coordinated changes across multiple cell types, and provide a molecular foundation for developing targeted therapies to prevent fibrosis progression and reduce cancer risk. The potential therapeutic targets identified in this study offer novel avenues for mechanistic studies on these pathways, which can be potential therapeutic targets, for the challenging pediatric CP disease entity.

## Data sharing

Sequencing data have been deposited in the Gene Expression Omnibus (GEO), accession no. GSE325443.

## Data Availability

All data produced in the present study are available upon reasonable request to the authors

https://www.ncbi.nlm.nih.gov/geo/query/acc.cgi?acc=GSE325443

## Acknowledgements

This research was performed with the support of the Network for Pancreatic Organ donors with Diabetes (nPOD; RRID:SCR_014641), a collaborative type 1 diabetes research project supported by Breakthrough T1D and The Leona M. & Harry B. Helmsley Charitable Trust (Grant#3-SRA-2023-1417-S-B). The content and views expressed are the responsibility of the authors and do not necessarily reflect the official view of nPOD. Organ Procurement Organizations (OPO) partnering with nPOD to provide research resources are listed at https://npod.org/for-partners/npod-partners/.

## Contributors

M.A.-E.-H., Y.W. conceived the study. M.A.-E.-H., Y.W., and F.A. designed the study. F.A. carried out experiments. X.X., Y.W., D.A., and F.A. performed data analyses. F.A., J.G., P.C., K.K., R.B., and C.W. contributed to sample collection. R.K. helped with library prep and sequencing. M.E.M.-F., and A.D. provided critical discussion and experimental suggestions. E.H.P. performed histological analyses. FA. and X.X. prepared manuscript with input from the coauthors. M.A.-E.-H., and Y.W. supervised the project. All authors provided critical feedback and helped shape the research, analysis and manuscript. All authors have reviewed and approved the final version of the manuscript.

## Supplementary material

**Supplementary Figure 1.**
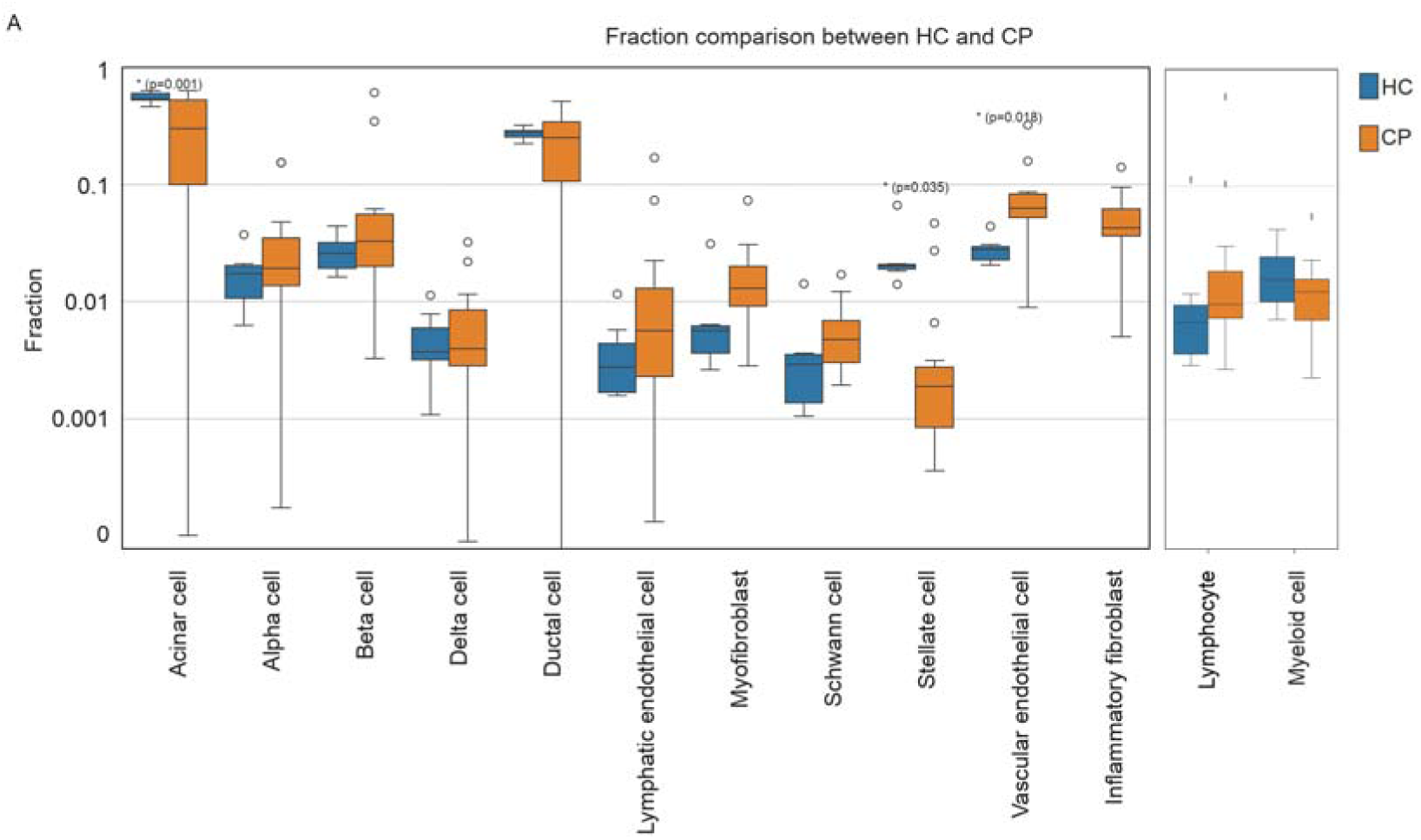
Cell-type fraction comparison between healthy control (HC) and chronic pancreatitis (CP). Box plots show the fractions of major pancreatic parenchymal, stromal, endothelial, and immune cell populations in HC (blue) and CP (orange) samples, including acinar, alpha, beta, delta, ductal, lymphatic endothelial, myofibroblast, Schwann, stellate, vascular endothelial, inflammatory fibroblast, lymphocyte, and myeloid cells. The y-axis represents the fraction of each cell type per sample on a log-scale. Each point denotes an individual sample. p-values are indicated above the corresponding cell types; with significant differences indicated by asterisks. Statistical significance was assessed using Welch’s two-sided unpaired t-test, and p-values ≤0.05 were considered statistically significant.

**Supplementary Figure 2.**
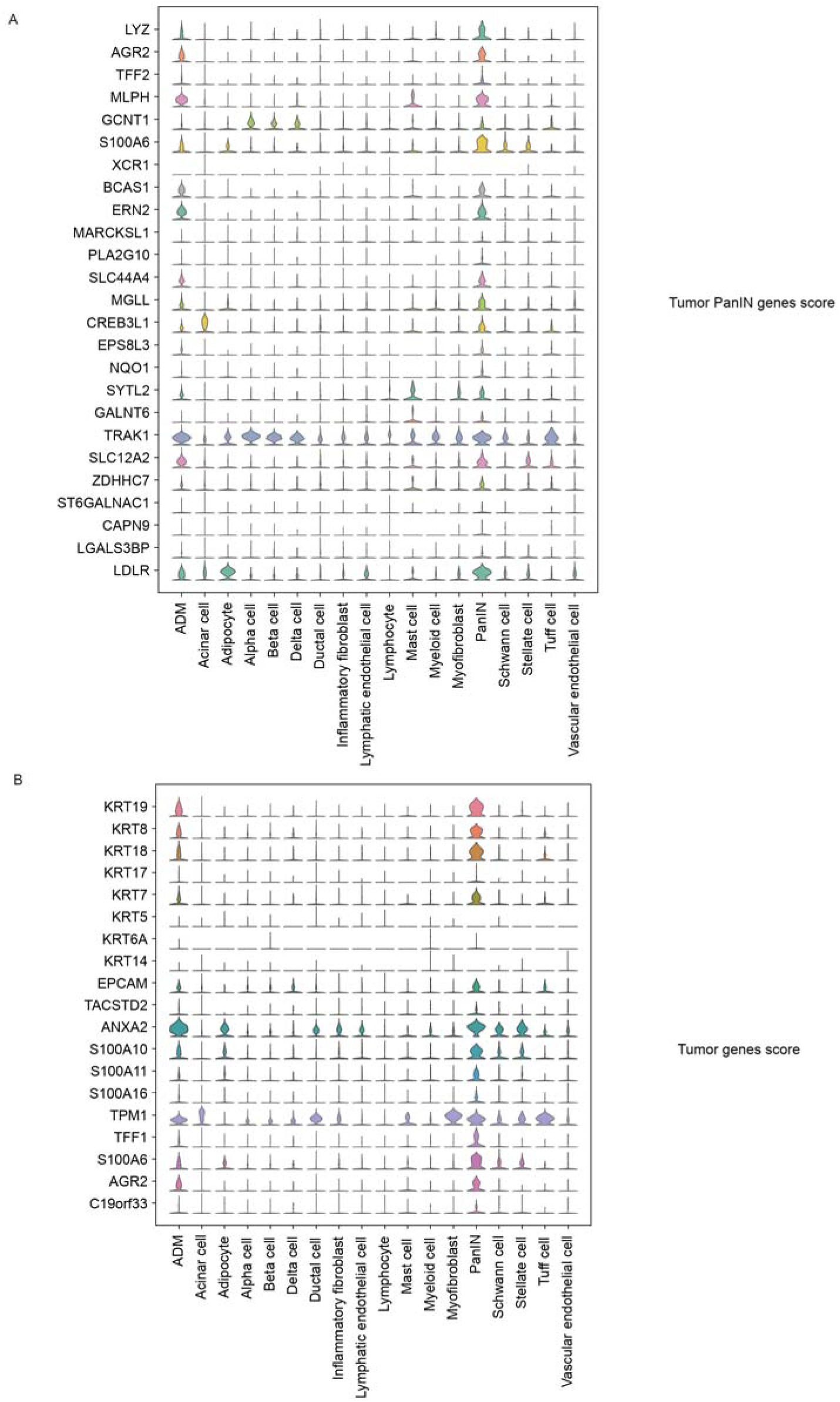
Expression of PanIN and tumor (PDAC) related gene signature in CP patients. Stacked violin plots illustrating the expression of PanIN-associated genes **(A)** and tumor-associated genes **(B)** across cell populations in CP samples, highlighting elevated expression in ADM and PanIN cells relative to other populations.

**Supplementary Figure 3.**
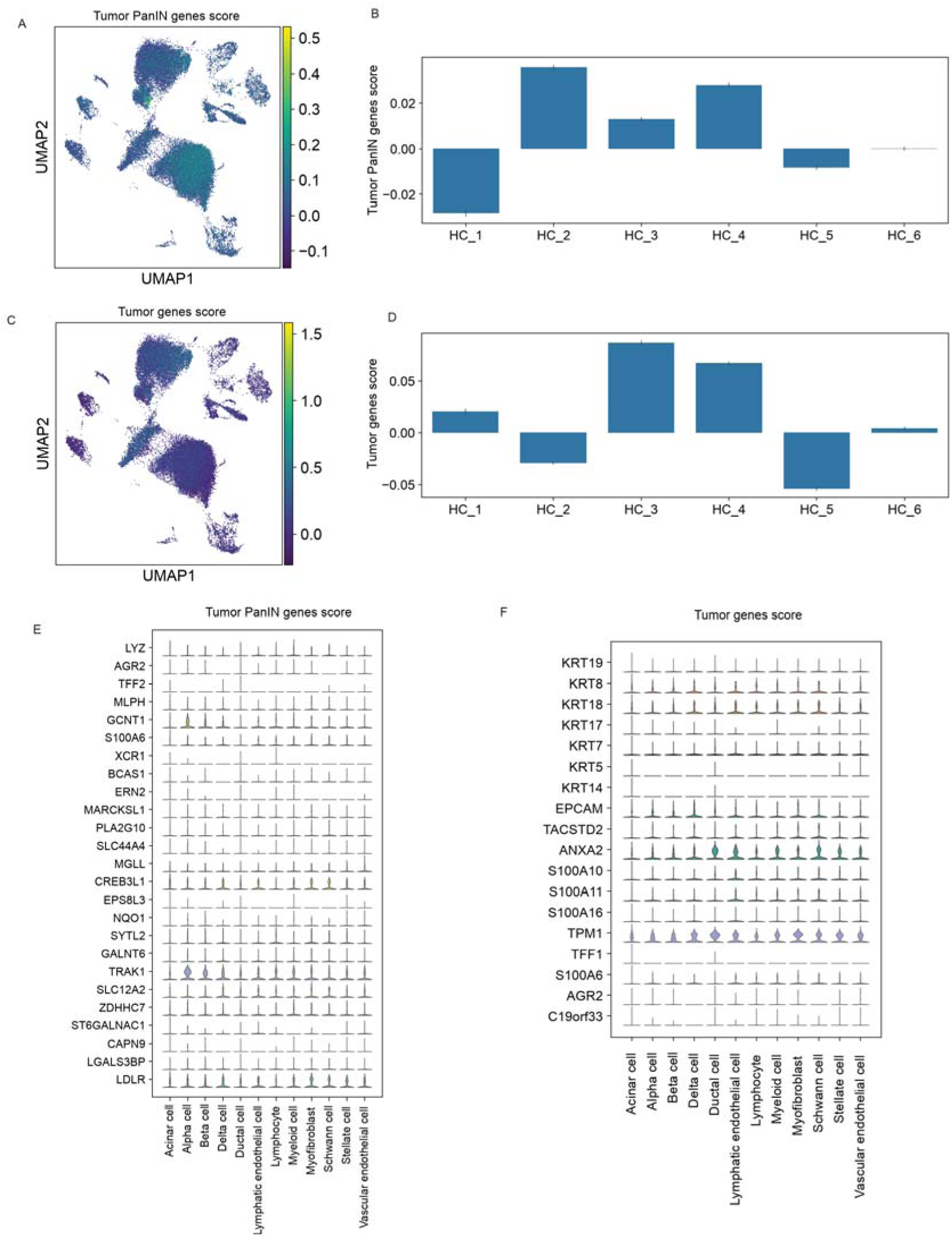
Tumor and PanIN gene signature scores in healthy control (HC) pancreatic cells. **(A)** UMAP projection of HC colored by the PanIN gene signature score. **(B)** Sample level quantification of the PanIN gene score across individual HC donors (HC_1-HC_6). **(C)** UMAP projection of HC cells colored by the tumor (PDAC) gene signature score. **(D)** Sample level tumor gene signature scores for individual HC donors (HC_1-HC_6). Stacked violin plots showing expression of representative PanIN-associated genes **(E)** and tumor-associated gene **(F)** across annotated HC cell types.

**Supplementary Figure 4.**
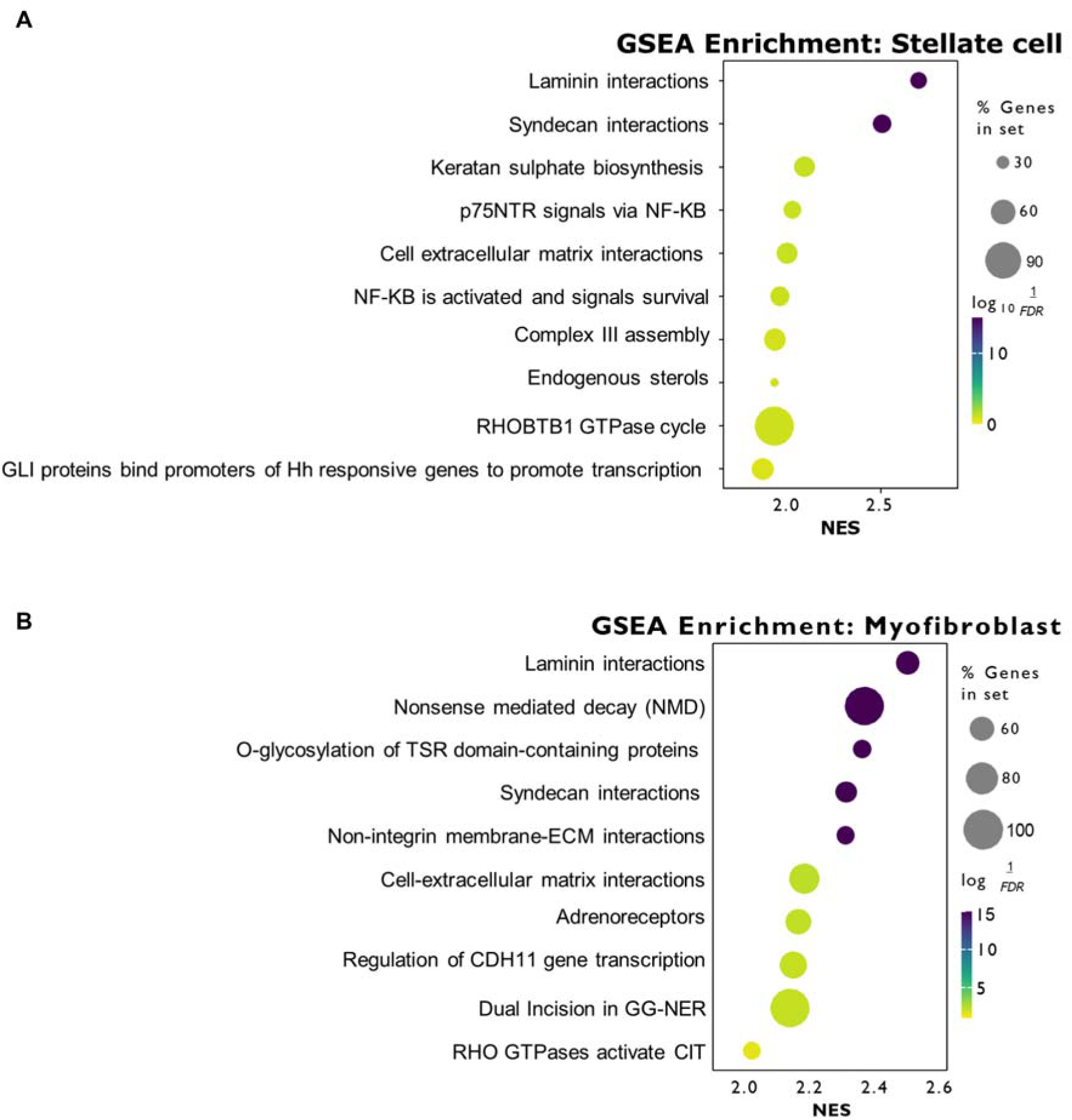
GSEA pathways in stellate cells and myofibroblasts. Dot plot showing Gene Set Enrichment Analysis (GSEA) of pathways enriched in stellate cell **(A)** and myofibroblasts **(B)** from CP tissues. Dot size indicates the percentage of genes in each pathway; color represents −log10(FDR), and the x-axis shows normalized enrichment score (NES). Top enriched pathway for both stellate cells and myofibroblasts was laminin interactions.

**Supplementary Table 1.** Comparison of fraction of cell type populations in CP and HC.

| Cell type | HC<br>(mean) | HC<br>(SD) | CP<br>(mean) | CP<br>(SD) | p-value |
| --- | --- | --- | --- | --- | --- |
| Acinar cell | 0.5793 | 0.0504 | 0.3015 | 0.2282 | 0.000354 |
| Alpha cell | 0.0169 | 0.0111 | 0.0379 | 0.0493 | 0.139478 |
| Beta cell | 0.0254 | 0.0104 | 0.093 | 0.167 | 0.140374 |
| Delta cell | 0.0039 | 0.0022 | 0.0075 | 0.0088 | 0.155463 |
| Ductal cell | 0.2799 | 0.0342 | 0.2372 | 0.168 | 0.361918 |
| Lymphatic<br>endothelial cell | 0.0038 | 0.0039 | 0.0219 | 0.045 | 0.143075 |
| Myofibroblast | 0.0047 | 0.0016 | 0.018 | 0.017 | 0.009086 |
| Schwann cell | 0.0023 | 0.0012 | 0.0061 | 0.0044 | 0.006745 |
| Stellate cell | 0.019 | 0.0026 | 0.0067 | 0.013 | 0.002926 |
| Vascular endothelial<br>cell | 0.0257 | 0.0041 | 0.0814 | 0.0762 | 0.013409 |
| Inflammatory<br>fibroblast | 0 | 0 | 0.0553 | 0.0336 | 0.000017 |

**Supplementary Table 2.** Histology scores (ADM, PanIN, Fibrosis) for H&E slides from CP pancreas tissues (n=13). Two samples were excluded due to unavailable tissue slides.

| Specimen ID | ADM score | PanIN score | Fibrosis score |
| --- | --- | --- | --- |
| CP_1 | Tissue N/A for histology |  |  |
| CP_2 | >75% | yes, frequent | 90% replacement; hyalinized |
| CP_3 | none | none | minimal |
| CP_4 | 50% | one small focus | 30 |
| CP_5 | 30% | few foci | 60 |
| CP_6 | none | one small focus | 10% |
| CP_7 | Tissue N/A for histology |  |  |
| CP_8 | none | none | 10%, hyalinized |
| CP_9 | 30% | focal | 75% replacement |
| CP_10 | none | none | fatty replacement |
| CP_11 | none | minimal (few foci) | 5% hyalinized |
| CP_12 | none | none | none |
| CP_13 | 10% | none | 10% |
| CP_14 | 20% | none | 10% |
| CP_15 | 10% | minimal (few foci) | 30% hyalinized |

